# Patterns and persistence of SARS-CoV-2 IgG antibodies in Chicago to monitor COVID-19 exposure

**DOI:** 10.1101/2020.11.17.20233452

**Authors:** Alexis R. Demonbreun, Thomas W. McDade, Lorenzo Pesce, Lauren A. Vaught, Nina L. Reiser, Elena Bogdanovic, Matthew P. Velez, Ryan R. Hsieh, Lacy M. Simons, Rana Saber, Daniel T. Ryan, Michael G. Ison, Judd F. Hultquist, John T. Wilkins, Richard T. D’Aquila, Brian Mustanski, Elizabeth M. McNally

**Affiliations:** Center for Genetic Medicine, Northwestern University Feinberg School of Medicine; Department of Pharmacology, Northwestern University Feinberg School of Medicine; Department of Anthropology and Institute for Policy Research, Northwestern University; Division of Cardiology, Department of Medicine, Northwestern University Feinberg School of Medicine; Division of Infectious Diseases, Department of Medicine, Northwestern University Feinberg School of Medicine; Institute for Sexual and Gender Minority Health and Wellbeing and Department of Medical Social Sciences, Northwestern University; Division of Organ Transplantation, Dept. of Surgery, Northwestern University Feinberg School of Medicine; Department of Preventive Medicine, Northwestern University Feinberg School of Medicine; Department of Biochemistry and Molecular Genetics, Northwestern University

**Keywords:** COVID-19, SARS-CoV-2, serological testing, IgG, ELISA, dried blood spots, essential worker, nucleocapsid, receptor binding domain

## Abstract

**Background:** Estimates of seroprevalence to SARS-CoV-2 vary widely and may influence vaccination response. We ascertained IgG levels across a single US metropolitan site, Chicago, from June 2020 through December 2020.

**Methods:** Participants (n=7935) were recruited through electronic advertising and received materials for a self-sampled dried blood spot assay through the mail or a minimal contact in person method. IgG to the receptor binding domain of SARS-CoV-2 was measured using an established highly sensitive and highly specific assay.

**Results:** Overall seroprevalence was 17.9%, with no significant difference between method of contact. Only 2.5% of participants reported having had a diagnosis of COVID-19 based on virus detection, consistent with a 7-fold greater exposure to SARS-CoV-2 measured by serology than detected by viral testing. The range of IgG level observed in seropositive participants from this community survey overlapped with the range of IgG levels associated with COVID-19 cases having a documented positive PCR positive test. From a subset of those who participated in repeat testing, half of seropositive individuals retained detectable antibodies for 3-4 months.

**Conclusions:** Quantitative IgG measurements with a highly specific and sensitive assay indicate more widespread exposure to SARS-CoV-2 than observed by viral testing. The range of IgG concentration produced from these asymptomatic exposures is similar to IgG levels occurring after documented non-hospitalized COVID-19, which is considerably lower than that produced from hospitalized COVID-19 cases. The differing ranges of IgG response, coupled with the rate of decay of antibodies, may influence response to subsequent viral exposure and vaccine.

**Graphical Abstract:** 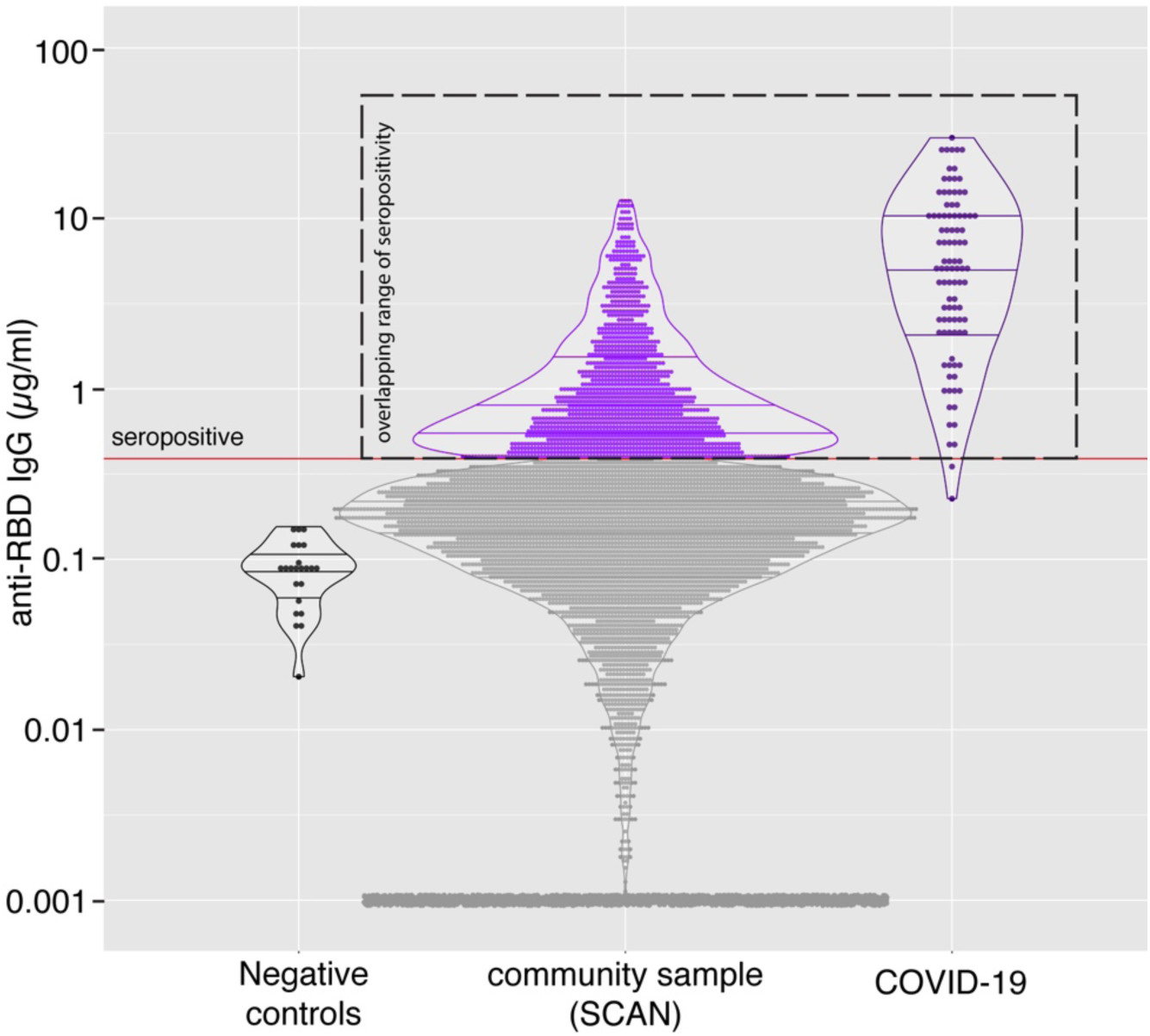

## Introduction

The presence of serum antibodies specific to SARS-CoV-2, the virus that causes COVID-19, reflects prior exposure. Seroprevalence, or seropositivity, estimates range from 3-50% depending on the population surveyed, method of testing, and viral antigen target (1-13). Most tests measure antibody content in blood or serum, which often necessitates contact with a health care facility. Point-of-care lateral flow devices have also been employed but lack both the sensitivity and specificity of a laboratory-performed measurement (3, 14).

As an alternative, dried blood spots (DBS) can be easily collected at home using a simple finger prick method, and the used in a laboratory performed assay to yield quantitative IgG measurements. We used DBS in an assay indexes IgG levels to an antibody with known affinity to yield a concentration in micrograms per ml (µg/ml). The antigen in this assay is restricted to the receptor binding domain (RBD), a small and specific domain within the SARS-CoV-2 spike protein; multiple studies have documented specificity of 97.7-100% when using this target, owing to RBD’s limited sequence homology to other viruses (1, 15, 16). DBS sampling was performed on nearly 8000 participants across Chicago. Approximately half of the participants were recruited from a wide socioeconomic range by zipcodes. These participants received and returned their DBS test kits through the United States Postal Service. The remaining participants were primarily non-healthcare-providing students, staff, and faculty affiliated with a medical school who received and returned DBS kits having minimal contact with the study staff.

The total cohort included a mix of individuals who were working outside the home as well as those working inside the home during the testing interval. The sampling began in late June 2020, in a period when local shelter-in-place orders were partially relaxed and continued through December 2020. Overall, the IgG seroprevalence was 17.9%, with similar seroprevalence among samples ascertained through mail and those obtained through onsite DBS kit distribution. Moreover, seroprevalence did not differ between those working inside or outside the home. The IgG range of seropositive individuals overlapped with documented, non-hospitalized COVID-19 cases consistent with widespread exposure to COVID-19 through the second half of 2020.

## Results

### Prevalence of IgG antibodies to the receptor binding domain of SARS-CoV-2

The Screening for Coronavirus Antibodies in Neighborhood (SCAN) study uses an at-home testing strategy to measure IgG antibodies to the receptor binding domain (RBD) of the SARS-CoV-2 spike protein (16). IgG assays with RBD as antigen have shown 97.7-100% specificity and 81-89% sensitivity (15-17). Participants received a kit to provide a dried blood spot sample (DBS) from a finger prick, and DBS cards were used in a quantitative laboratory-performed ELISA. Between June and December 2020, 7935 SCAN samples were collected. Of SCAN participants, 195 (2.5%) reported having COVID-19 from a prior, positive diagnostic test for SARS-CoV-2 virus. Participants (n=5898) were recruited advertising and social media, receiving and returning test materials through the mail (no-contact method) (**Figure 1**). An additional 2037 were solicited through email and in-person contact to provide and retrieve the DBS materials (contact method). Seropositivity in SCAN participants utilizing the no-contact method was 18.2 % (n=1072 of 5898), while the seropositivity among the group who used the contact method was 17.3 % (n=352 of 2037) (odds ratio 1.06; p= 0.4; CI 0.93-1.23).

**Figure 1.**
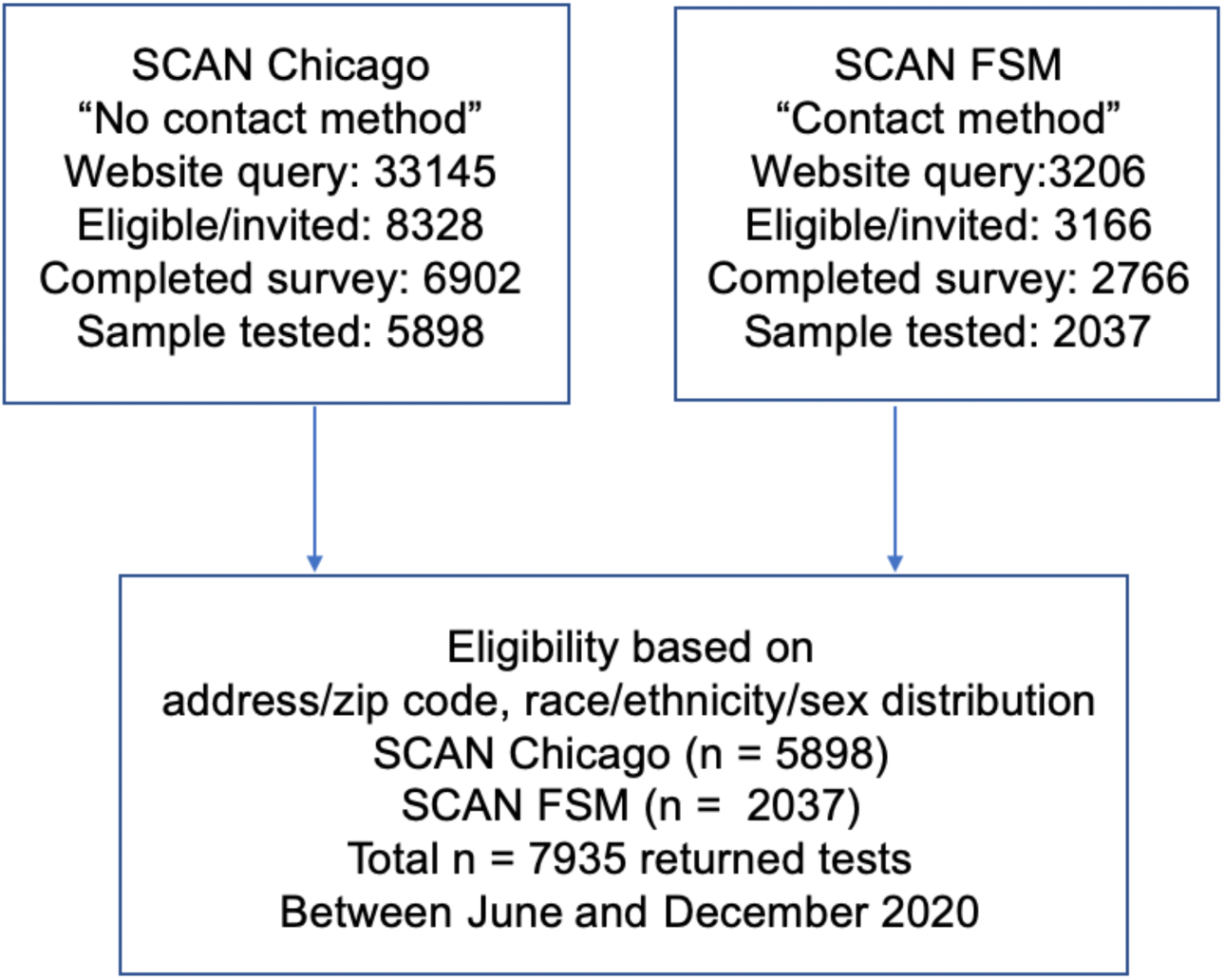
Flow diagram for recruitment into Screening for Coronavirus Antibodies in Neighborhoods (SCAN) studies. Participants were recruited to enter queries to the SCAN website through social media, news coverage and paid advertising with focus on zip codes throughout Chicago. Individuals were screened for eligibility based on living in specific zip codes and recruited to promote a racially/ethnically mixed cohort with adequate representation of males and females, and then invited to complete a health questionnaire survey. Dried blood spot kits were sent to all those eligible who completed the survey. These participants received and returned dried blood spot kits through mail (no contact method) with an 85% return rate. A second cohort was recruited by email through the Northwestern’s Feinberg School of Medicine (FSM), and these individuals received blood spot kits in person and returned kits on site (contact method) with a 74% return rate.

Of the total 7935 SCAN participants, 195 (2.5%) reported having COVID-19 with a prior positive virus test, with 169 of 195 (86.6%) seropositive for RBD IgG. Of these 7935 participants, 1424 (17.9%) were seropositive and 6511(82.1%) were seronegative. This represents 7 times more seropositive samples than confirmed SARS-CoV-2 nucleic acid positivity in the SCAN cohort. Seropositivity was similar between males 18.8% (615 of 3278) and females 17.4% (809 of 4657) (**Table 1**). Seropositivity by age group varied slightly from 20.9% (18-29 yrs), 17.2 % (30-39 yrs), 17.6% (40-49 yrs), 18.0% (50-59 yrs), and 14.0% (60+ yrs) (**Table 2**).

**Table 1.** Seropositivity by birth sex.

| n= 7935 | Seropositive |  | Seronegative |  | Total |  |
| --- | --- | --- | --- | --- | --- | --- |
|  | n | % | n | % | n | % |
| Male | 615 | 18.8 | 2663 | 81.2 | 3278 | 100.0 |
| Female | 809 | 17.4 | 3848 | 82.6 | 4657 | 100.0 |

**Table 2.** SCAN results by age.

| SCAN n=<br>7935 | Seropositive |  | Seronegative |  | Total |  |
| --- | --- | --- | --- | --- | --- | --- |
|  | n | % | n | % | n | % |
| 18-29 | 411 | 20.9 | 1560 | 79.1 | 1971 | 100.0 |
| 30-39 | 439 | 17.2 | 2110 | 82.8 | 2549 | 100.0 |
| 40-49 | 273 | 17.6 | 1281 | 82.4 | 1554 | 100.0 |
| 50-59 | 182 | 18.0 | 829 | 82.0 | 1011 | 100.0 |
| 60+ | 119 | 14.0 | 731 | 86.0 | 850 | 100.0 |

### IgG serum levels in SCAN overlap with IgG levels in outpatient COVID-19 cases

The CR3022 antibody has known affinity for the target antigen, making it possible to quantify IgG directed at RBD. There was no difference in the mean IgG level in seronegative samples compared to samples collected in 2018 (pre-COVID-19 samples), (median 0.09 µg/ml vs 0.09 µg/ml). The median IgG concentration from seropositive samples from SCAN was 0.75 µg/ml. As a comparison, the median IgG from those with a COVID-19 diagnosis based on virus detection (n=96), but who did not require hospitalization, was 5.2 µg/ml, while the median IgG from those requiring ICU hospitalization was 98.5 µg/ml (**Figure 2**). While the median levels differ between documented COVID-19 cases and SCAN seropositive participants, the range of IgG showed wide overlap between these groups indicating extensive range in IgG response among those with outpatient COVID-19 and those with asymptomatic exposure.

**Figure 2.**
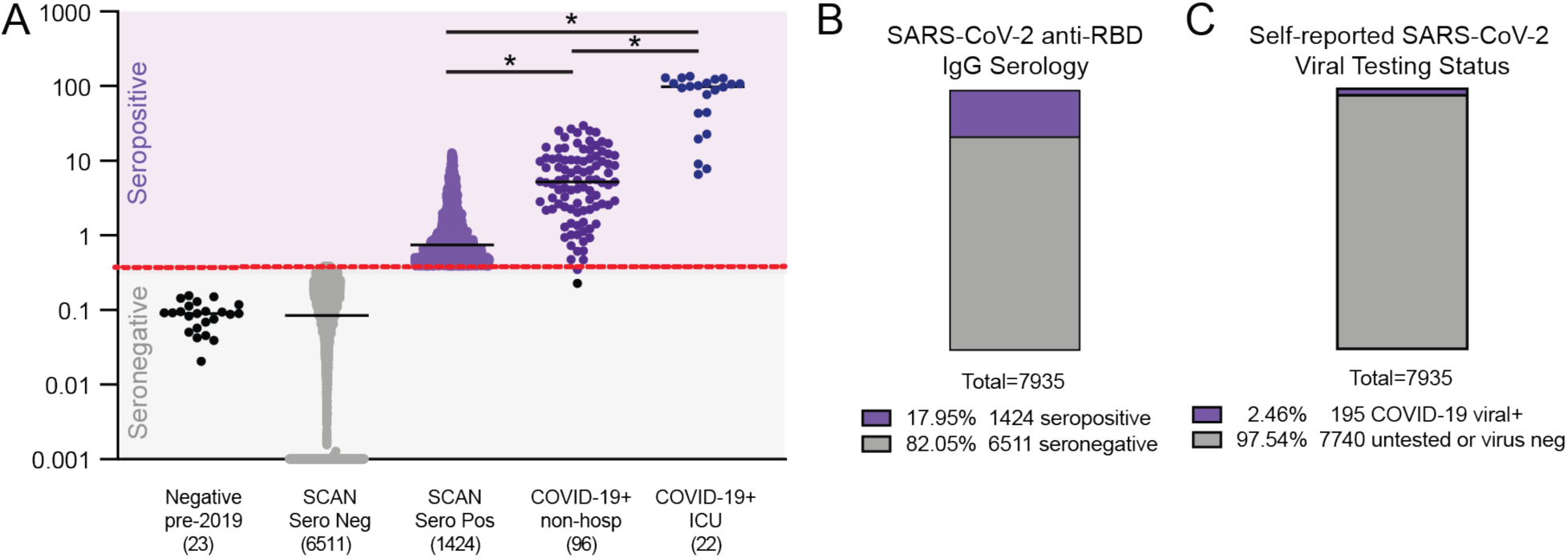
Quantitative measure of IgG directed to the receptor binding domain (RBD) of SARS-CoV-2 spike glycoprotein. Samples were acquired through the Screening for Coronavirus Antibodies in Neighborhood (SCAN) between June 2020 and December 2020 (n= 7935). **A**) Samples acquired before 2019 constituted the Negative, Pre-COVID group (leftmost column, black dots, median 0.09μg/ml), and the mean IgG was similar to the SCAN seronegative group (second column, gray dots, median 0.09 µg/ml). The median IgG range in SCAN seropositive samples (middle column, light purple) was 0.75 µg/ml. The median IgG range from outpatient, non-hospitalized COVID-19 samples (dark purple, 4^th^ column) was 5.2μg/ml. The range between SCAN seropositive and outpatient COVID-19 showed significant overlap. Both SCAN seropositive and outpatient COVID-19 cases had much lower IgG levels than ICU hospitalized COVID-19 cases (median 98.5 µg/ml). The SARS-CoV-2 RBD IgG ELISA seropositive threshold is marked by the red line at 0.39µg/ml as validated previously (16). Comparing seropositive groups * p<0.0001 by Wilcoxon-Mann-Whitney Test. Both true negatives and the SCAN seronegative groups were significantly different than all seropositive groups. **B**) 17.95% (1424 of 7935) of SCAN samples were seropositive. **C**) 2.46% (195 of 7935) reported having a positive SARS-CoV-2 viral diagnostic test.

### Similar levels of SARS-CoV-2 RBD IgG antibodies in essential and non-essential workers

Between March 21 and May 30, 2020 the state of Illinois and the city of Chicago were under a shelter-in-place order, except for certain workers and essential trips. Between June and August 2020, these orders were gradually relaxed but limits on gatherings of more than 50 people and restrictions on indoor dining/bars remained in effect. In late November 2020, a stay-at-home advisory was put in place in the City of Chicago asking residents to only leave home for essential activities, reflecting the earlier second wave experienced in the Midwest. SCAN participants reported whether they left their place of residence and interacted with others at the workplace during the shelter-in-place or stay-at home intervals. Essential (n=2829) and non-essential (n=5106) groups had similar percent seropositivity at 18.4% and 17.7%, respectively (**Figure 3A and 3B**). The two groups were well matched by design for age, sex, and race (**Table 3**). The median RBD IgG level and distribution was not different (p=0.86) between the seropositive essential (n =520, median 0.75 µg/ml) and seropositive non-essential groups (n = 904, median 0.74 µg/ml) (**Figure 3C**).

**Table 3.** Demographics of essential (working outside the home) and non-essential (working from home) personnel.

| SCAN n= 7935 | Non-Essential |  | Essential |  | Total |  |
| --- | --- | --- | --- | --- | --- | --- |
|  | n | % | n | % | n | % |
| Birth Sex |  |  |  |  |  |  |
| Male | 2156 | 42.2 | 1122 | 39.7 | 3278 | 41.3 |
| Female | 2950 | 57.8 | 1707 | 60.3 | 4657 | 58.7 |
| Race/Ethnicity |  |  |  |  |  |  |
| NH Asian | 886 | 17.4 | 507 | 17.9 | 1393 | 17.6 |
| NH Black | 394 | 7.7 | 178 | 6.3 | 572 | 7.2 |
| Hispanic/Latinx | 1093 | 21.4 | 653 | 23.1 | 1746 | 22.0 |
| NH White | 2454 | 48.1 | 1357 | 48.0 | 3811 | 48.0 |
| NH Multiracial | 212 | 4.2 | 90 | 3.2 | 302 | 3.8 |
| NH Other | 67 | 1.3 | 44 | 1.6 | 111 | 1.4 |
| PCR Status |  |  |  |  |  |  |
| Negative/Unknown | 4970 | 97.3 | 2747 | 97.1 | 7717 | 97.3 |
| Positive | 121 | 2.4 | 74 | 2.6 | 195 | 2.5 |
| Missing | 15 | 0.3 | 8 | 0.3 | 23 | 0.3 |
| IgG Antibody Result |  |  |  |  |  |  |
| Seronegative | 4202 | 82.3 | 2309 | 81.6 | 6511 | 82.1 |
| Seropositive | 904 | 17.7 | 520 | 18.4 | 1424 | 17.9 |
|  | Median | IQR | Median | IQR | Median | IQR |
| Age | 38.0 | 20.0 | 36.0 | 17.0 | 37.0 | 18.0 |

**Figure 3.**
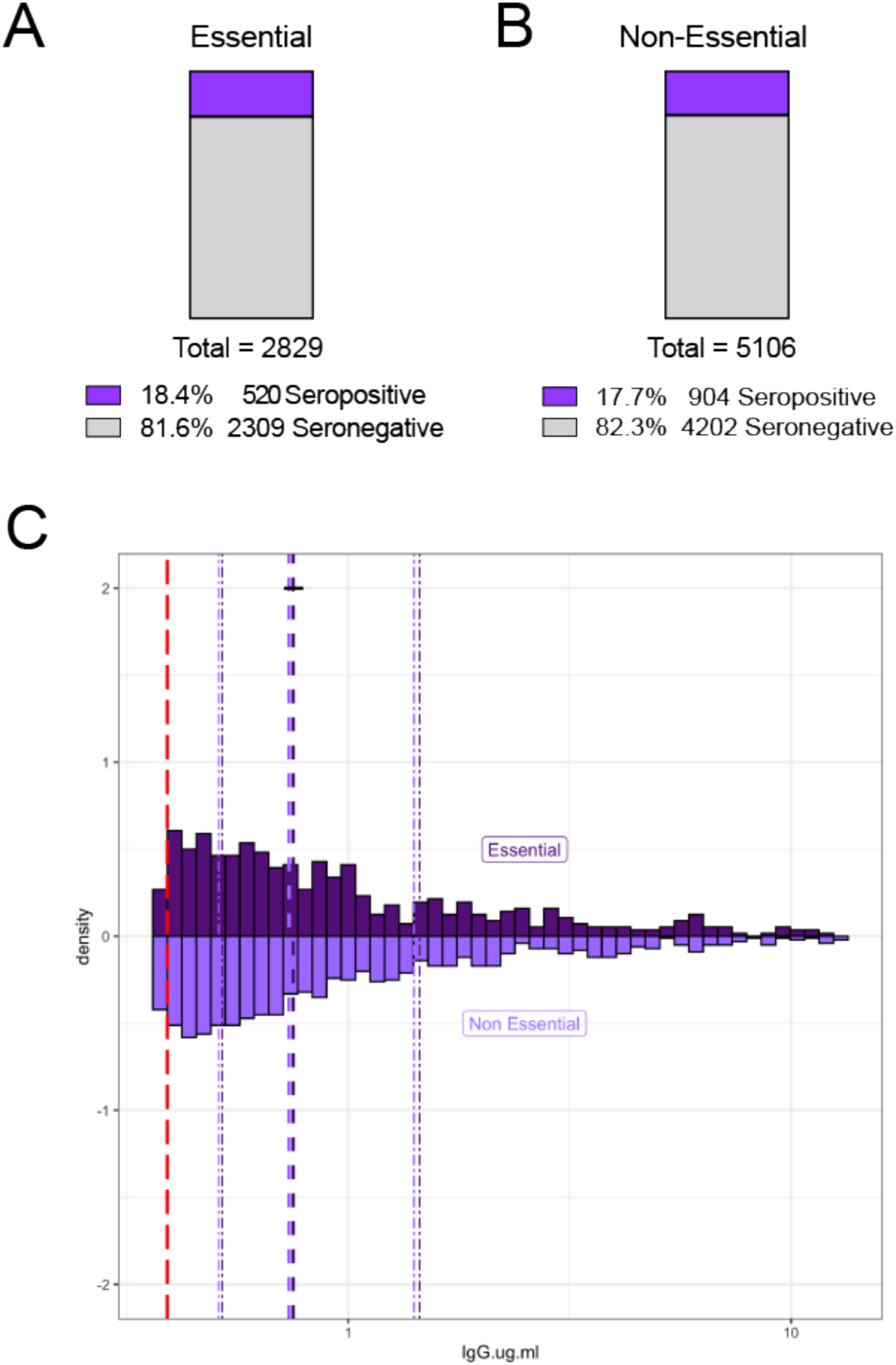
Similar rates of SARS-CoV-2 RBD IgG seropositivity between those who reported working outside the home (“essential”) and those working from home (“non-essential”). 7935 unique SCAN community-acquired samples were acquired between June and December 2020 from the Chicago area. Participants self-reported whether they left their residence for work (essential) and interacted with co-workers / public. **A and B)** Essential and non-essential reported groups have similar percent seropositivity at 18.4 % and 17.7%, respectively. **C**) Essential (n=520) and non-essential (n=904) groups had similar distributions of SARS-CoV-2 RBD IgG seropositivity with a median of 0.75 µg/ml and 0.74 µg/ml, respectively. The SARS-CoV-2 RBD IgG ELISA positivity threshold is denoted with the red dotted line at 0.39 µg/ml. Dashed purple lines represent quartiles. (Two-sample Kolmogorov-Smirnov test p=0.86.)

### Only modest agreement between RBD and nucleocapsid serology status

We assayed 28 samples from individuals who recovered from symptomatic COVID-19 with positive virus diagnostic testing, and 92 suspected samples but who did not reporting a positive virus diagnostic test result. Agreement between RBD IgG and nucleocapsid classification was modest for both the COVID-19-positive (K=0.20; 95% CI: 0.00-0.59) and suspected SCAN group (K=0.21; 95% CI: 0.12-0.32) groups (**Figure 4**). Specifically, 6 of 28 (21.4%) COVID-19-positive had no IgG to nucleocapsid using a hospital-performed, FDA authorized assay. Only one sample of these 28 was below the limit of detection in the RBD assay, and this same sample was also negative in the nucleocapsid assay. The remaining 27 of 28 were positive for RBD IgG. A similar analysis of 92 suspected positive SCAN samples was conducted; this cohort was selected to skew towards seropositivity based on reported possible exposure or mild symptoms. Of the 92 “suspected” samples, 65 (70.7%) samples had IgG to RBD. Of these 65 positive samples, 20 samples had IgG to both RBD and nucleocapsid, while the remaining 45 had only IgG to RBD and not to nucleocapsid. In both groups, there were no samples that were RBD negative and nucleocapsid positive. These data suggest limited sensitivity of nucleocapsid testing.

**Figure 4.**
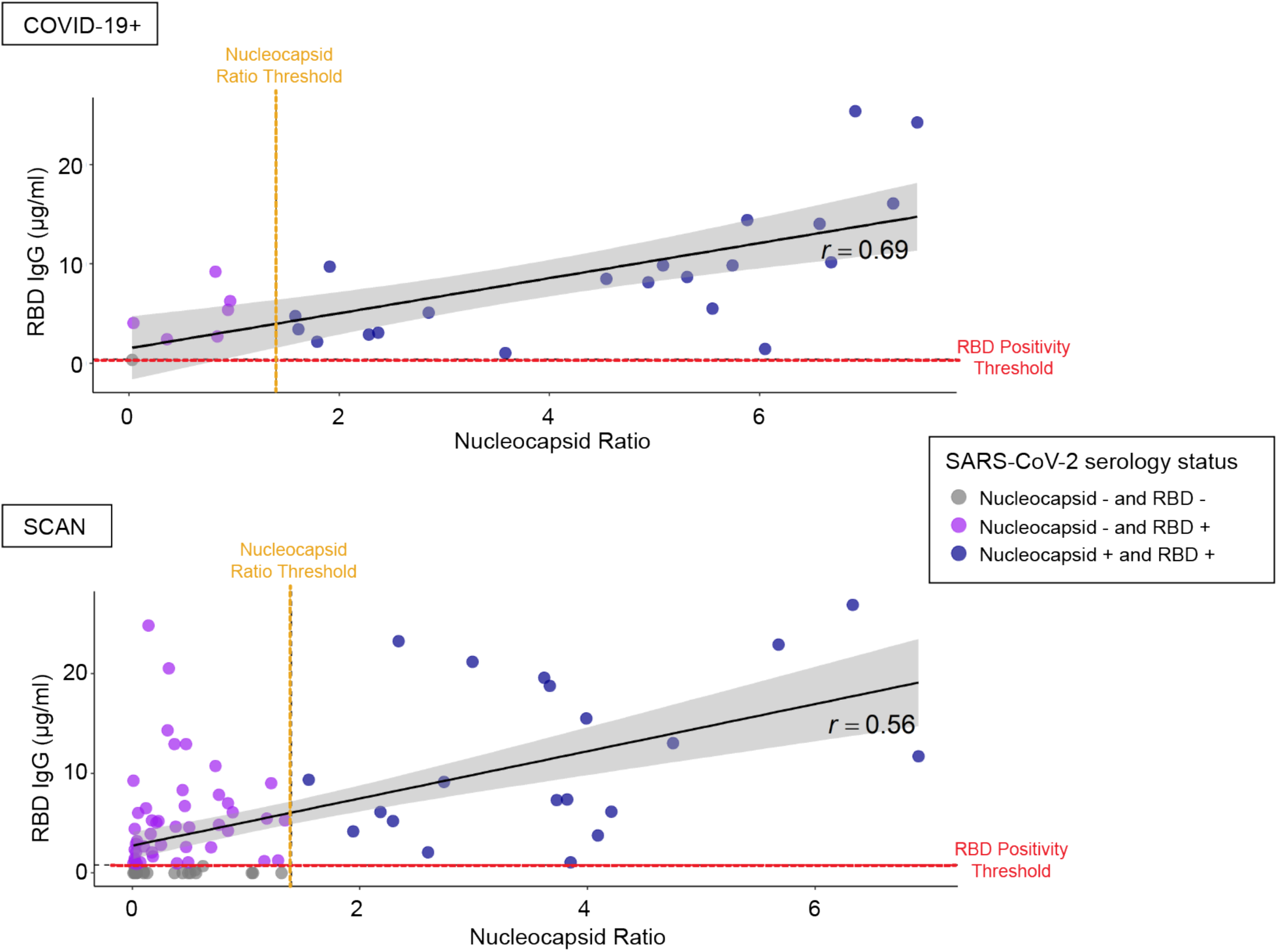
Modest agreement between SARS-CoV-2 RBD IgG and nucleocapsid seropositivity. Twenty-eight COVID-19+ viral positive, non-hospitalized samples and 92 SCAN samples with untested/negative COVID-19 status were analyzed for the presence of SARS-CoV-2 RBD IgG and nucleocapsid IgG antibodies using a hospital performed test. Six of 28 (21.5%) known COVID-19+ viral positive samples were nucleocapsid negative but were RBD IgG positive. One of 28 COVID-19+ viral positive samples was seronegative on both platforms. Of the 92 unknown COVID-19 status samples, 20 (21.7%) samples were both nucleocapsid and RBD IgG positive, while 45 (48.9%) samples were nucleocapsid negative and RBD IgG positive. Agreement between RBD IgG and nucleocapsid classification was modest for both the known COVID-19 viral positive (K=0.20; 95% CI: 0.00-0.59) and unknown COVID-19 status (K=0.21; 95% CI: 0.12-0.32) samples. The black diagonal line and gray shaded area represent the simple linear regression of RBD IgG on nucleocapsid ratio and the 95% confidence interval band respectively. The SARS-CoV-2 RBD IgG ELISA positivity threshold is denoted with the red dotted line at 0.39 µg/ml. The SARS-CoV-2 nucleocapsid positivity threshold is denoted with the orange dotted line at ratio 1.4.

### Persistence of SARS-CoV2 RBD IgG antibodies over time

We monitored change in RBD IgG concentration across the entire cohort of seropositive SCAN participants (n= 1305) over time. IgG concentration in a cross-sectional comparison at each time point remained similar over 26 weeks from June to December 2020 (**Figure 5A**). We separately assessed longevity of IgG persistence in 87 seropositive participants with repeat sampling 3-4 months (mean 103 days) from their first detected seropositivity (**Figure 5B**). The median IgG concentration at day 0, the first day of seropositivity, was 0.59 µg/ml, above the 0.39 µg/ml positivity threshold. After 84-132 days the median IgG concentration decreased to 0.397 µg/ml (p= 0.45). Forty-four of 87 seropositive samples (50.6%) remained seropositive over 84-132 days after the first seropositive result. Of those remaining seropositive, 24 of 44 (54.5%) had similar or increased RBD IgG levels upon resampling, while 20 of 44 (45.5%) had decreased RBD IgG concentration. These data show that half of SCAN participants who were seropositive at their first test, had detectable IgG antibodies against SARS-CoV-2 RBD at 3-4 months.

**Figure 5.**
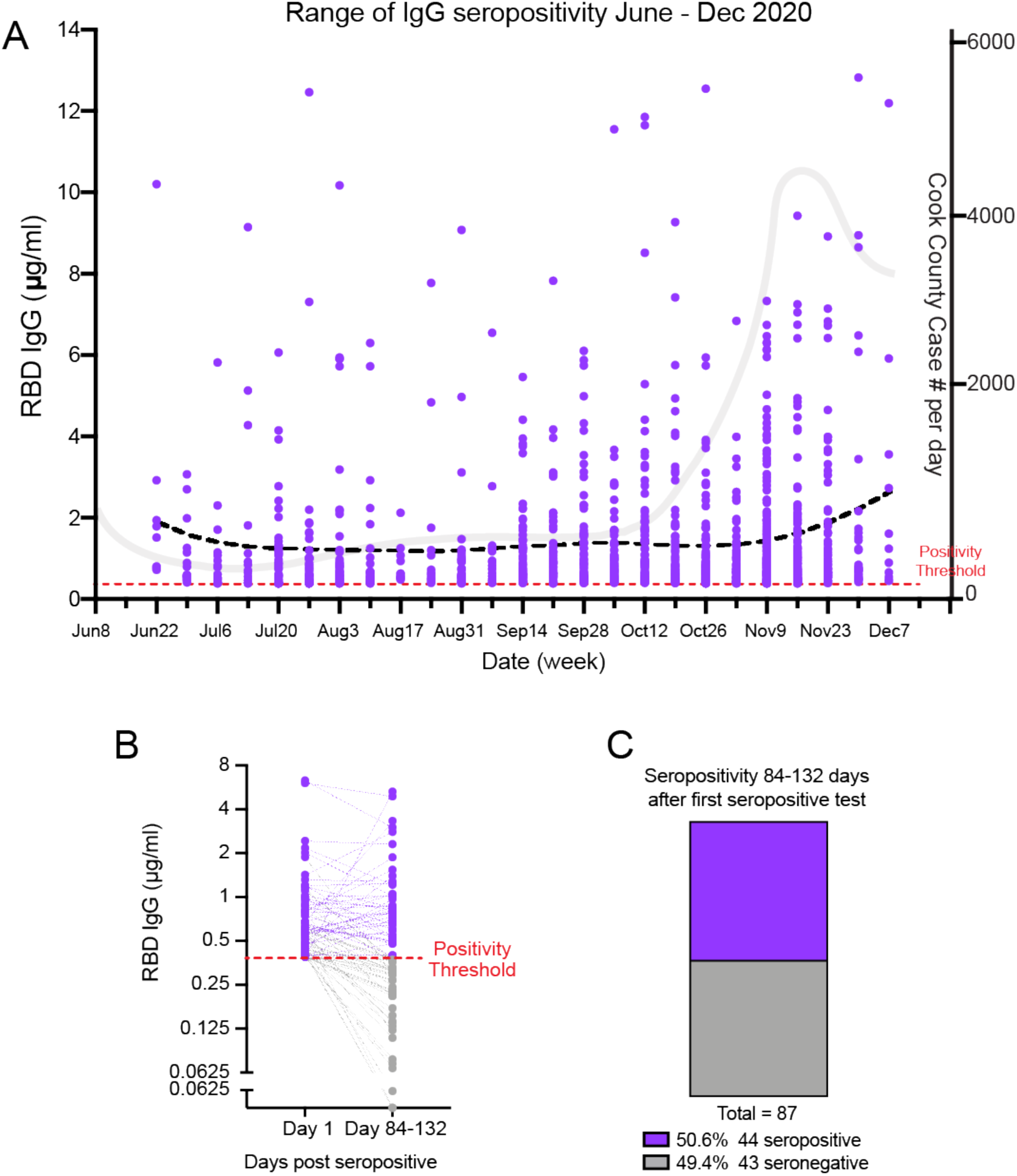
Detectable SARS-CoV-2 RBD IgG antibodies after 3-4 months in SCAN community samples. **A**) Two hundred and eighty-six seropositive RBD IgG SCAN sample concentrations plotted as function of calendar week of acquisition. Lowess curve (black dotted line) is steady across 26 weeks of sampling. Grey line illustrates COVID-case number per day in Cook County. **B and C**) Eighty-seven seropositive SCAN participants (purple dots) were resampled 84 – 132 days post the first seropositive RBD IgG test (mean 103 days). Dotted lines connect the same participant over time. Eighteen of 87 (50.6%) samples remained seropositive after 3-4 months with 43 (49.4%) samples converting to seronegative (grey dots). Time points were not significantly different p = 0.45.

Three individuals with stable seropositive RBD IgG level, over at least a three month period, reported subsequently having a positive SARS-CoV-2 diagnostic virus test result despite the presence of antibodies to RBD. Each reported having COVID-19 symptoms, and none required hospitalization. The participants sought testing for COVID-19 after experiencing several days of cough, or altered taste and smell. RBD IgG levels 14-28 days after the positive SARS-CoV-2 diagnostic test were significantly increased in all three individuals (**Figure 6**). This seroprevalence pattern is likely to reflect re-exposure to SARS-CoV-2, confirmed by a viral diagnostic result at the second exposure, and demonstrates a strong immune response after a second exposure.

**Figure 6.**
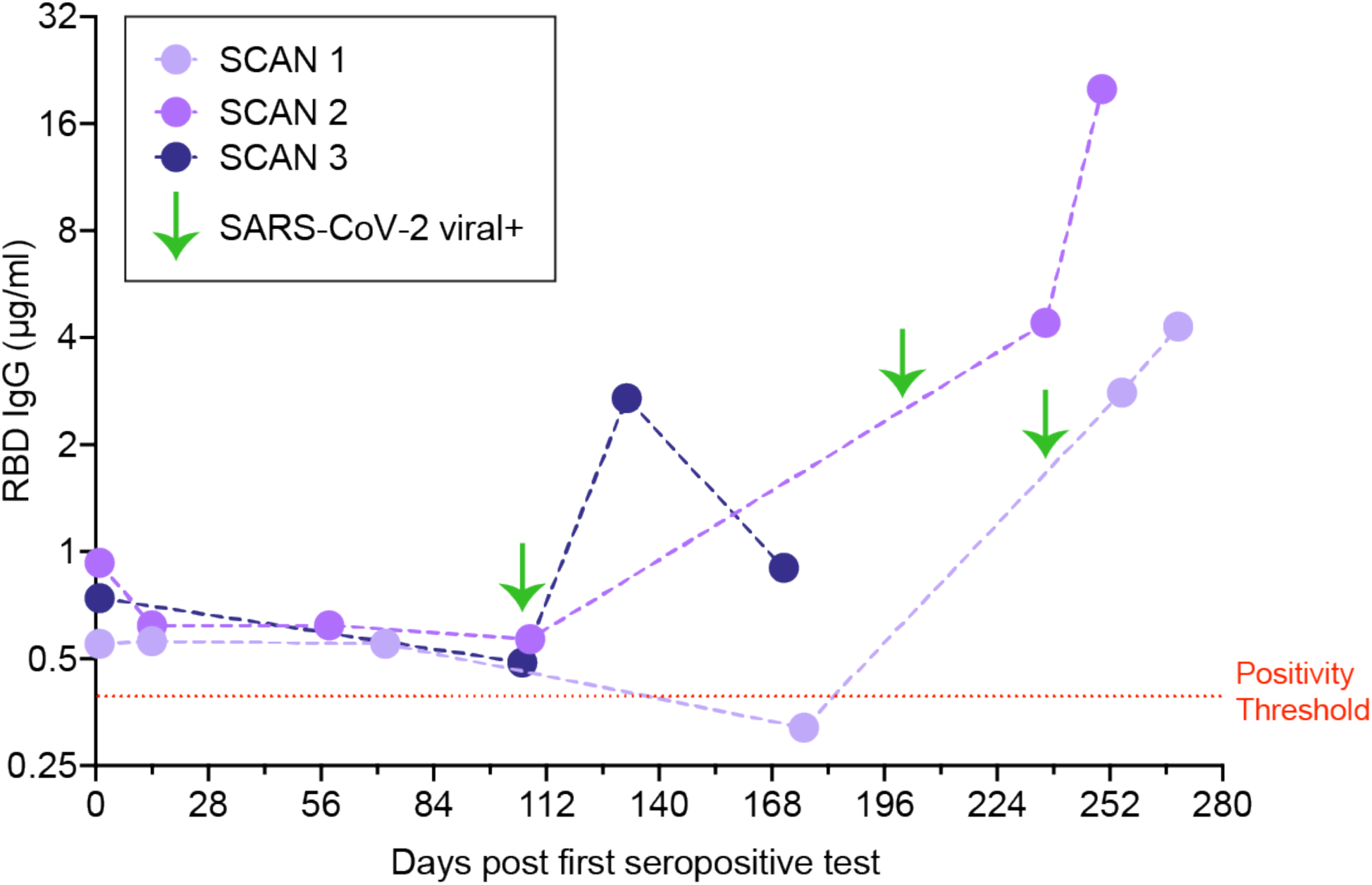
Re-exposure to SARS-CoV-2. Three IgG RBD seropositive individuals (SCAN 1, 2, 3) were observed to have a marked increase in IgG to RBD 14-28 days after positive SARS-CoV-2 diagnostic viral testing (green arrows). Mild symptoms were noted including cough, altered taste and smell. None required hospitalization.

## Discussion

### Estimating SARS-CoV-2 exposure with quantitative determination of RBD antibodies

We surveyed the presence of IgG to a restricted region of the spike glycoprotein since this domain has been shown to be highly specific to SARS-CoV-2 (17). The 17.9% seropositivity rate in this study is consistent with this antibody test identifying 7-fold greater exposure than noted with virus diagnostic testing, supporting previous predictions (18). The median level of IgG among seropositive cases was lower than documented COVID-19 outpatient cases, but the range of IgG overlapped between these two groups. RBD IgG levels were much higher in ICU hospitalized COVID-19 cases. It is the RBD portion of the spike protein that mediates viral entry into cells, and some antibodies to the RBD can be neutralizing and protect against cellular invasion. Whether the RBD antibodies detected in the SCAN study and mildly symptomatic or asymptomatic COVID-19 are neutralizing or protective in nature is unknown. It also remains to be determined whether or how much these lower levels of RBD IgG serve to prime the immune system on re-exposure or for subsequent vaccination.

### RBD versus nucleocapsid antibodies

We found that among individuals with documented, symptomatic COVID-19, there was a lower detection rate for the nucleocapsid antigen, using a hospital performed, FDA authorized test. The discordance between anti-nucleocapsid and anti-RBD IgG antibody detection was noted in a study of COVID-19-positive serum using other independent platforms (Pearson’s correlation of 0.65) (19). Among RBD IgG seropositive individuals that were asymptomatic or mildly symptomatic, there was even less concordance between anti-nucleocapsid and anti-RBD IgG antibody detection. Gudbjartsson et al reported similar findings in an Islandic cohort of persons untested or negative for SARS-CoV-2, where only 44.3% of anti-RBD IgG positive samples were also positive for anti-nucleocapsid IgG antibody (5). This differential sensitivity may represent differences in antibody response, testing platform, or both. Like others, we favor the use of RBD IgG determination since this affords high specificity given the unique nature of the target domain.

Testing for COVID-19 virus became much more widely available beginning in May and June 2020 and increased exponentially thereafter, yet only 2.5% of individuals in this survey reported a positive COVID-19 test consistent with a high rate of asymptomatic exposure to virus across many areas of Chicago. Although the SCAN cohort derives from a wide range of Chicago neighborhoods, the cohort could be biased by those self-enrolling due to higher concerns for SARS-CoV-2 exposure risk.

### Persistence of IgG directed to RBD

During the summer of 2020, local restrictions imposed during spring 2020 were eased. This included allowing retail stores to open, outdoor dining, permitting larger gatherings, and gym reopening. Mitigation measures were then reinstated in November 2020 related to the early second wave that spread across the Midwest and Chicago during October. We observed a similar seroprevalence among participants who self-reported as leaving the home for work during the restricted periods and those who remained at home. We observed three cases of documented SARS-CoV-2 viral infection in individuals with prior seropositivity, and each developed a strong “boost” of IgG to RBD after having documented viral infection. We expect that this pattern reflects what occurs with second exposure and potentially mimicks what might occur with vaccination. The SCAN platform, which relies on simple at home monitoring combined with laboratory precision, is positioned to help address this, and other, knowledge gaps.

## Methods

### Study Recruitment

Participants were recruited through two mechanisms. Community-based participants were recruited from ten zip codes in Chicago through social media advertising and news articles. Alternatively, staff, students and faculty from the Northwestern University Feinberg School of Medicine in Chicago IL were sent an email describing the study with a link to the website. Participants were screened for eligibility (zip code and demographics or affiliation to Northwestern). Eligible participants were then invited to complete a questionnaire regarding health status, including COVID-19 symptoms. Community participants received materials for DBS collection through the USPS and returned their test kits using prepaid USPS envelopes provided to them by the study team. Those affiliated with Northwestern were given a specific time to collect DBS kits in person and were instructed to return their completed kits to a secure, unmanned collection box. Sample collection occurred between June and December 2020. Eighty-seven participants were resampled 84-132 days after the first seropositive RBD IgG test. Samples from 2018 (n=23) were used as negative controls. Comparator samples derived from 40 COVID-19 non-hospitalized cases, 22 hospitalized cases, and 110 samples from a healthcare worker study selected from a larger cohort of 1790 samples (20). These 110 samples were selected based on a high expected probability of concordance and discordance between the nucleocapsid IgG and RBD assays. For example, the sample was enriched for participants specifically who were nucleocapsid IgG+ and PCR+ (concordance) and for participants with COVID-19 symptoms and exposures who were nucleocapsid IgG negative (discordance). Thirty SARS-CoV2 positive samples were used from a study starting April 24, 2020 as comparators and two seropositive participants were resampled (16).

### Serological assay

The enzyme linked immunosorbent assay (ELISA) protocol was previously described (16, 17). As in the prior assay, samples were run in duplicate and reported as the average. Results were normalized to the CR3022 antibody with known affinity (Creative Biolabs #MRO-1214LC) (21). Participant sample anti-RBD IgG concentration (µg/ml) was calculated from the 4PL regression of the CR3022 calibration curve. A value >0.39µg/ml CR3022 was considered positive.

### Statistical Analysis

Statistical analyses were performed with Prism (GraphPad, La Jolla, CA) or R 4.2 (The R Foundation for Statistical Computing, http://www.R-project.org). The difference between groups used Wilcoxon-Mann-Whitney Test or the Kruskal-Wallis test if more than one group was tested. Two-sample Kolmogorov-Smirnov test was used to compare essential vs non-essential samples. Changes over time were depicted with first order locally weighted regression and smoothing (Lowess model). An unpaired two-tailed t-test was used to compare two groups, where appropriate. Pearson’s chi-squared test statistic was used to compare proportions.

## Data Availability

Written requests for data should be made by qualified researchers trained in human subject confidentiality protocols to the corresponding author.

## Study approvals

All research activities were implemented under protocols approved by the institutional review board at Northwestern University (#STU00206652, #STU00212371, #STU00212457, #STU00212472, and STU00212515).

## Author Contributions

LV, NR, MV, EB, RH processed samples and performed ELISAs.AD, LP, RS, DR managed the data, analyzed data, and generated figures. TM, EM, BM, JW, JH, and MI secured IRB approval and collected samples. AD, BM, TM, RD, EM provided critical input in study design and wrote the manuscript. All authors reviewed and approved the final version of the manuscript.

## Acknowledgments

Supported by NSF 2035114, NIH 3UL1TR001422-06S4, and a generous gift from Dr. Andrew Senyei and Noni Senyei.This work was supported by supplements to the NIH National Center for Advancing Translational Sciences grant UL1 TR001422 and UL1 TR002389 (JFH), the generous support of the Dixon Family Foundation (JFH), a supplement to the Northwestern University Cancer Center P30 CA060553 (JFH), and through a generous contribution from the Walder Foundation’s Chicago Coronavirus Assessment Network (JFH). We acknowledge the efforts of Grace Gallo in sample collection. The funding sources had no role in the study design, data collection, analysis, interpretation, or writing of the report.

